# Food Hygiene and associated factors among street food vendors of selected districts of Bagamati Province, Nepal: A cross-sectional study

**DOI:** 10.64898/2026.10.09.26365118

**Authors:** Safal Poudel, Aashma Dahal, Aishwarya Nangia

**Author notes:** Email: safal.

## Abstract

**Background:** Street food vending is a major source of food supply in Nepal. Poor food hygiene practices among street food vendors pose a health risk by increasing the risk of foodborne diseases. Studies regarding food hygiene knowledge and practices are needed to improve public awareness. Therefore, this study aimed to assess food hygiene knowledge and practice among street food vendors and their associated factors in selected districts of Bagamati Province.

**Methods:** A community-based cross-sectional analytical study was carried out among 186 street food vendors of Districts located in central Nepal. Face-to-face interviews were conducted using a semi-structured questionnaire and an observation checklist. Knowledge and practice levels were categorized as good and poor. Chi-square test, Fisher’s exact test, odds ratio, and Mann-Whitney U test were applied for data analysis.

**Results:** Less than half (39.8%) of the vendors had good food hygiene knowledge, while 91.4% of the respondents reported good food safety and hygiene practices. The education level of street food vendors was significantly associated with food hygiene knowledge (p = 0.001) and food safety and hygiene practices (p = 0.042). Significant association was not seen between overall food hygiene knowledge and overall food safety and hygiene practices levels.

**Conclusion:** The study identified a gap between food hygiene knowledge and self-reported food hygiene practices among street food vendors. Education, infrastructure, and regulations among street food vendors play a key role in good food hygiene practices more than knowledge alone. Policymakers should focus on training of food vendors, regular supervision, and monitoring of vending sites to enhance food safety.

## Background

Food may become contaminated during the production-to-consumption process through the introduction of microorganisms via water, air, dust, insects, rodents, and food vendors (Thapa et al., 2024). The measures and conditions that must be taken to ensure food safety from production to consumption are known as food hygiene. (1)

Globally, food market has increased significantly due to urbanization, industrialization, the growth of the tourism sector, and economic globalization. (2) With increasing urbanization and acculturation, street food vending is becoming popular around the world.(3) Due to urbanization, working people have a greater demand for ready-to-eat food because of less time to prepare their meals at home (4). Similarly, street food is cheaper in comparison to food from restaurants and is easy for people to buy and get delivered to their doorsteps too. (5) Around 2.5 billion people worldwide are estimated to consume street food every day Street foods are a major source of food for most people as they are easily accessible, affordable and have unique flavors. (6)

Street food vendors are those individuals who sell food without having a permanent structure Street food vendors are categorized as mobile vendors and static vendors. Mobile dealers use their heads, carts, bicycles, motorcycles, or tricycles to transport cooked and packaged food for resale from one location to another. Food is cooked, kept, and served to consumers at the stationary fixed stalls of static vendors. (7)These food vendors mostly operate in streets, car parks, factories, markets, and schools. (8)

The World Health Organization (WHO) defines foodborne illness as diseases either infectious or toxic, caused by pathogens that enter the digestive system through the ingestion of food Vendors usually operate under unhygienic conditions, are not licensed, not supervised, lack training in food safety and hygiene, and are unknown to the causes of foodborne disease. (9,10)Poor knowledge and practice on food preparation and handling driving to poor food hygiene. Furthermore, street food vending is an informal business that is not usually regulated. More than 200 diseases are caused by eating unsafe food with dangerous germs, viruses, parasites, or chemicals. (11) Each year more than 1 in 3 people of developing nations are affected by foodborne diseases. (12)

Overall, street vendors, who share an abundant market of food, are unaware about food hygiene and safety and despite economic contribution, street food vendors still lack food hygiene knowledge and skill. (13)To assess the knowledge, sanitary conditions and practice relating to food hygiene and safety among street food vendors becomes imperative in regard. Thus, this study aims to study these practices among street vendors quantitatively to inform the public and policies about the status of food safety.

## Methodology

The cross-sectional analytical study design was used for this study. For this study, six municipalities (three from Chitwan District and three from Makawanpur District) were selected. The chosen municipalities are as follows:

| Name of districts | Name of municipalities |
| --- | --- |
| Makawanpur | Hetauda-Sub Metropolitan City |
|  | Manahari Rural Municipality |
|  | Bakaiya Rural Municipality |
| Chitwan | Ichchhyakamana Rural Municipality |
|  | Ratnanagar Municipality |
|  | Kalika Municipality |

The study population for this study were street food vendors of selected areas of Chitwan and Makawanpur Districts. Street food vendors from selected areas and above 18 years of age. Food vendors below 18 years of age. Vendors selling only pre-packaged foods/drinks.

**Fig 1:**
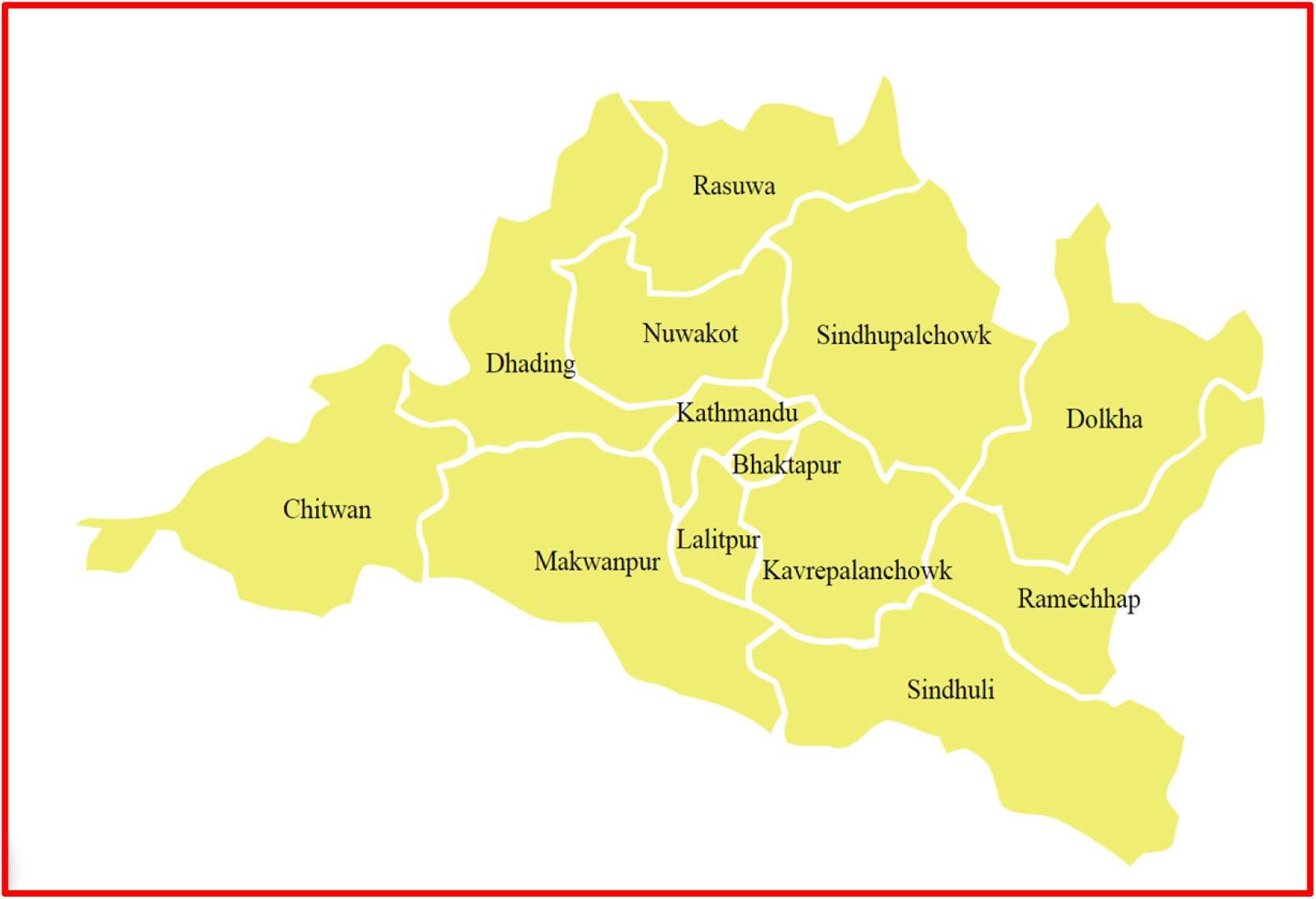
Map of Bagmati Province(14)

A multi-stage sampling technique was employed for the study. Two districts (Chitwan and Makawanpur) were selected conveniently in the first stage. In the second stage, three municipalities from each district (a total of six municipalities) were selected randomly and treated as clusters and in the final stage, study units in the given cluster were selected conveniently till the sample size was reached.

**Figure 2:**
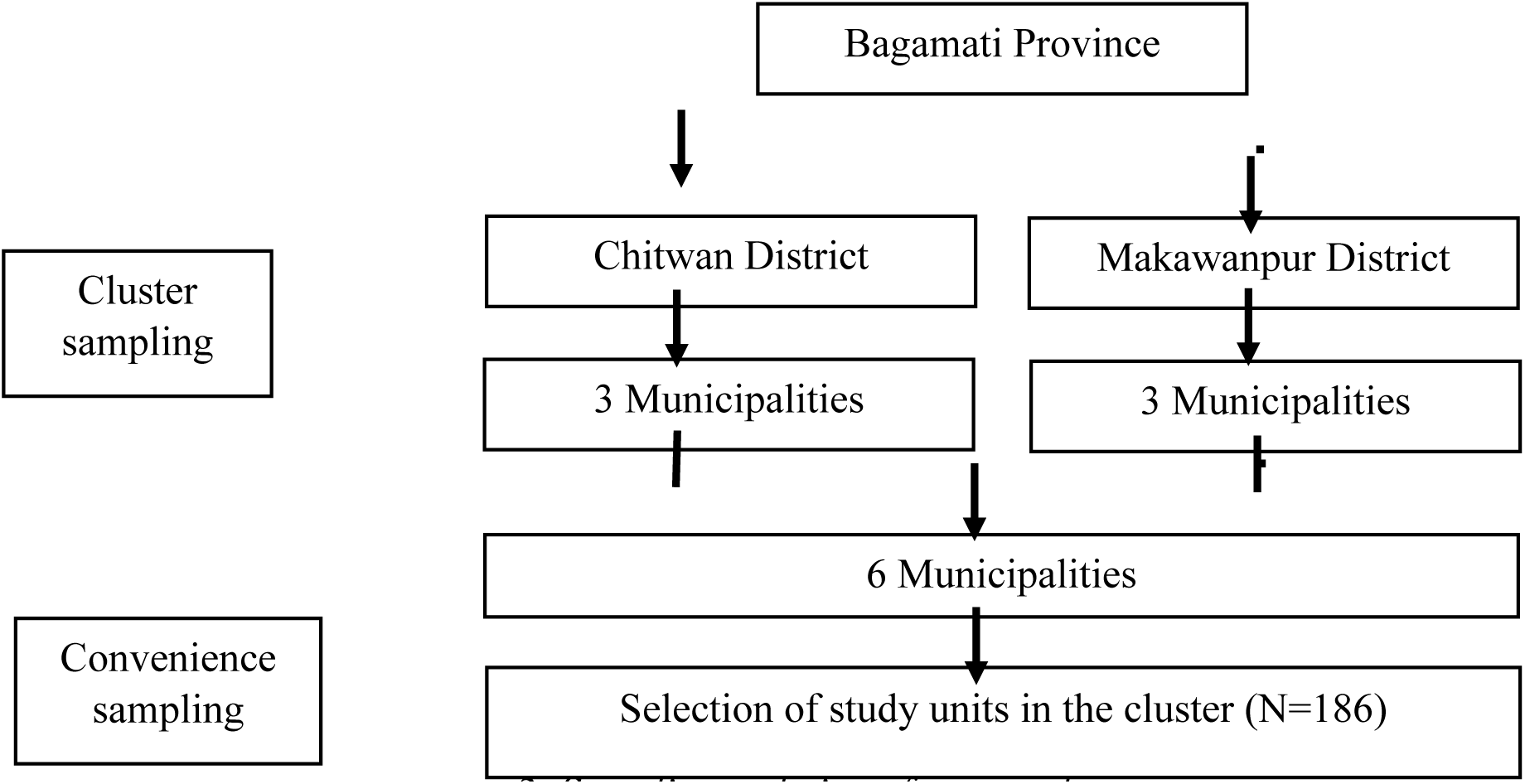
Sampling technique framework.

The sample size was calculated by using Cochran’s formula, n_0_= Z^2^pq/d^2^ with the precision of 5% (% confidence limits at an allowable error of 5%), p = 91.4% estimated proportion of sample size, where practice was good about food hygiene, we get 121. (13)Again, applying the 1.5 design effect of the sample for cluster sampling is: 121 * 1.5 = 181.5 (equals 182). But, to ensure an equal distribution across the six data collection sites, the sample per site was 31 (rounded up from 30.33), which resulted in the final total sample size of 186.

### 2.7 Data Collection tools and Techniques

Data were collected through face-to-face interviews by using semi-structured questionnaires, and an observation checklist was used for data triangulation. The tool was developed by the researcher based on extensive literature review, and discussion with the experts. The questionnaires were adopted from. (3,8,15) The study tool consists of four sections. Section I: Socio-demographic Information, Section II: Knowledge about food safety and hygiene, Section III: Food safety and hygiene practices of the vending unit and section IV was an observational checklist in order to validate the interview findings. The validity of the tool was maintained by continuous literature review of related research as well as consulting with experts. The instrument was pretested in 10% of the sample size. Pretesting was done in a similar population (*Gadi Municipality*) with samilar characteristics to check the clarity, feasibility and practicability of the study.

### Data Processing and Analysis

Coding and editing were done to prevent default and duplication. Excel and Statistical Package for the Social Sciences (SPSSv20) was used for data entry and analysis respectively. For quality control, daily screening of filled questionnaires was done to check completeness and consistency.

### 2.15 Ethical Consideration

Ethical clearance was obtained from IRC of Madan Bhandari Academy of Health Sciences.(IRC number: 102-82-83) Approval was also taken from all respective municipalities before data collection. Data collection started from 12 Dec 2025 till 15 Feb 2026. The purpose and objectives of the study were explained to the respondents and written consent was taken from each participant. Respondents were explained that their participation in the study was voluntary and they could withdraw from study any time. Confidentiality was maintained by using the given information only for the study purpose. Respondents were included without any discrimination regarding ethnicity, religion, age.

## Results

**Table 1:**
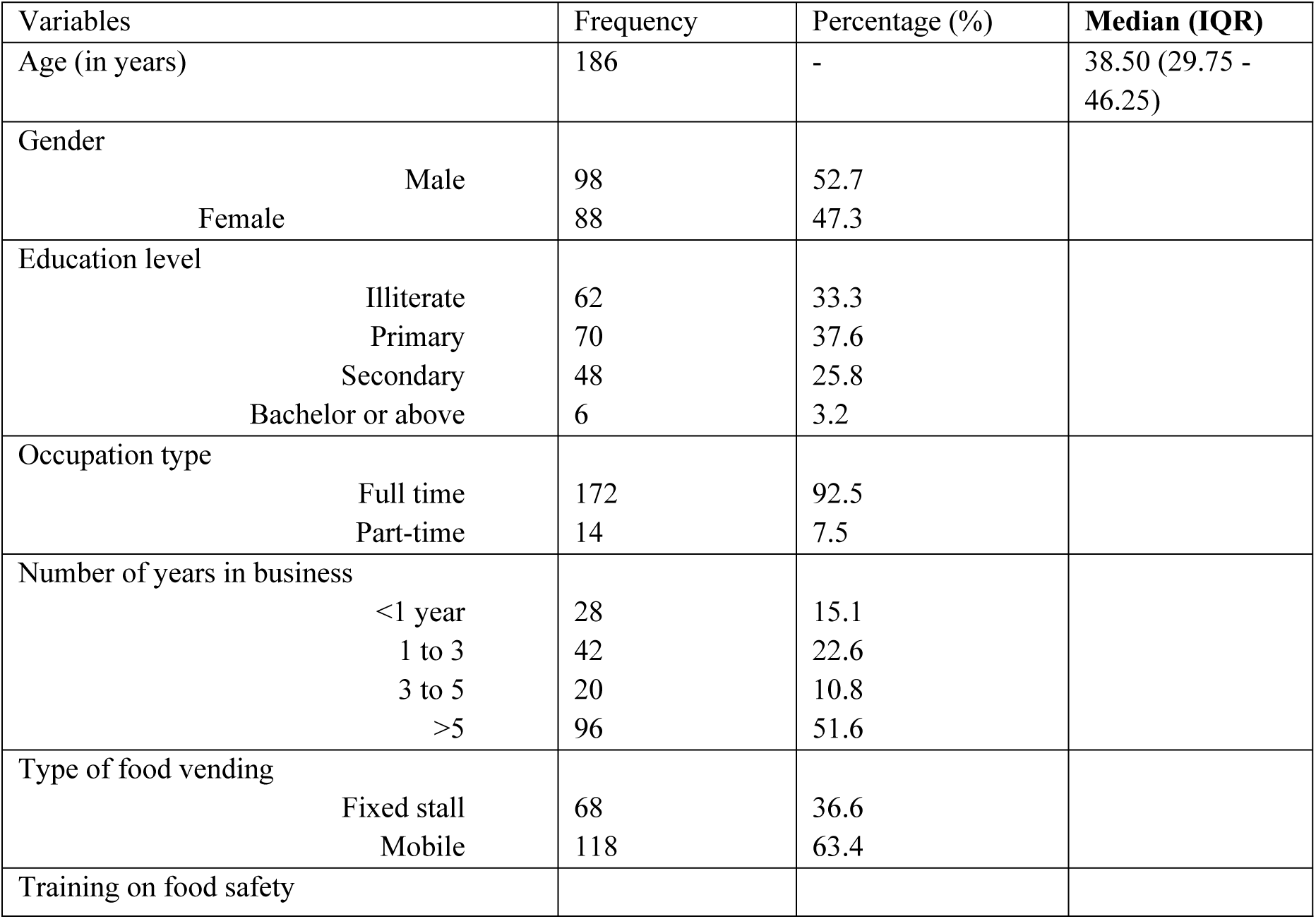
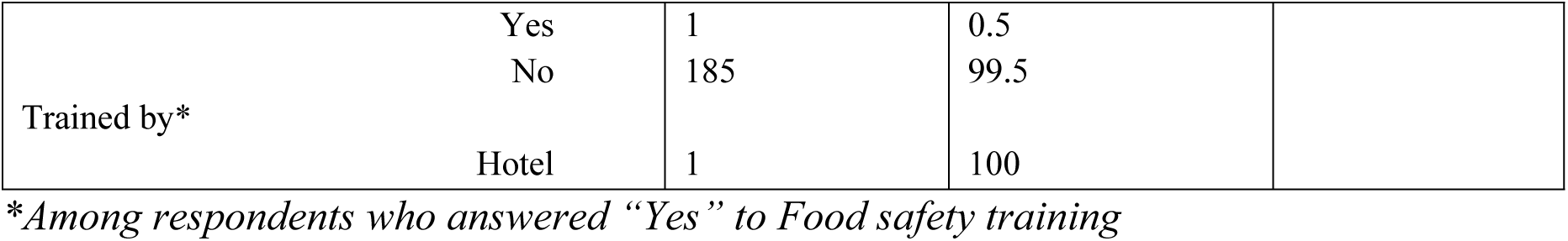
Socio-demographic characteristics of the participants(N=186)

**Tabel 2:** Food Hygiene Knowledge Among Food vendors.

| Characteristics | Frequency | Percentage (%) |
| --- | --- | --- |
| Know about Food Hygiene |  |  |
| Yes | 74 | 39.8 |
| No | 112 | 60.2 |
| If Yes, Definition of Food Hygiene* (n=74) |  |  |
| Clean, well-cooked food (Correct answer) | 69 | 93.2 |
| Other answer | 5 | 6.8 |
| Knowledge of symptoms/diseases caused by poor hygiene* |  |  |
| Yes | 70 | 94.6 |
| No | 4 | 5.4 |
| If “Yes” ** (Multiple response) (n=70), |  |  |
| Diarrhea | 58 | 82.9 |
| Nausea/Vomiting | 33 | 47.1 |
| Abdominal Pain | 40 | 57.1 |
| Others | 35 | 50 |
| Health problem affects food safety* (n=74) |  |  |
| yes | 67 | 90.5 |
| No | 7 | 9.5 |
| Knowledge level |  |  |
| Good Knowledge (>50%) | 73 | 39.2 |
| Poor Knowledge (<=50%) | 113 | 60.8 |

\*Among respondents who answered “Yes” to Knowledge on food hygiene.
\*\*Among respondents who answered “Yes” to Knowledge of symptoms/diseases caused by poor hygiene.

**Table 3:**
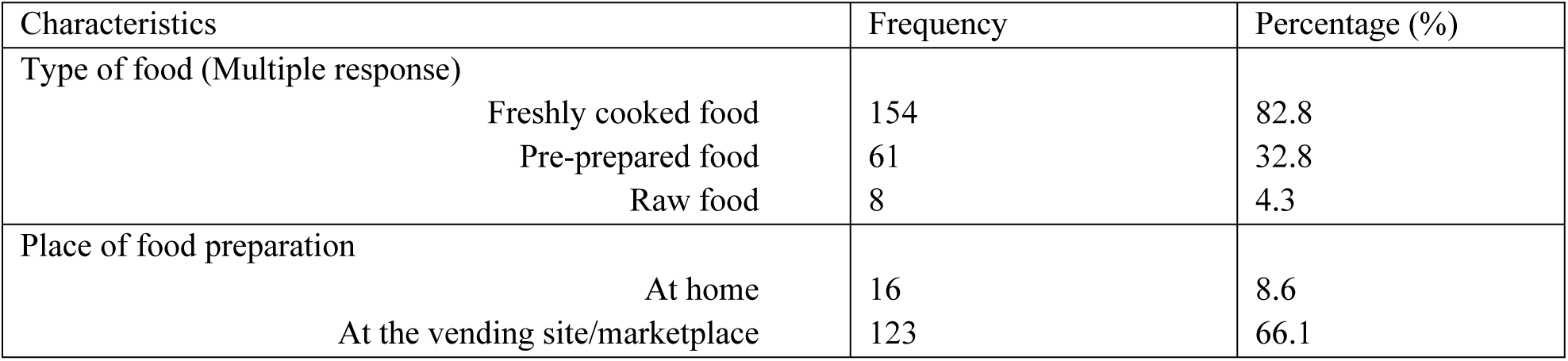
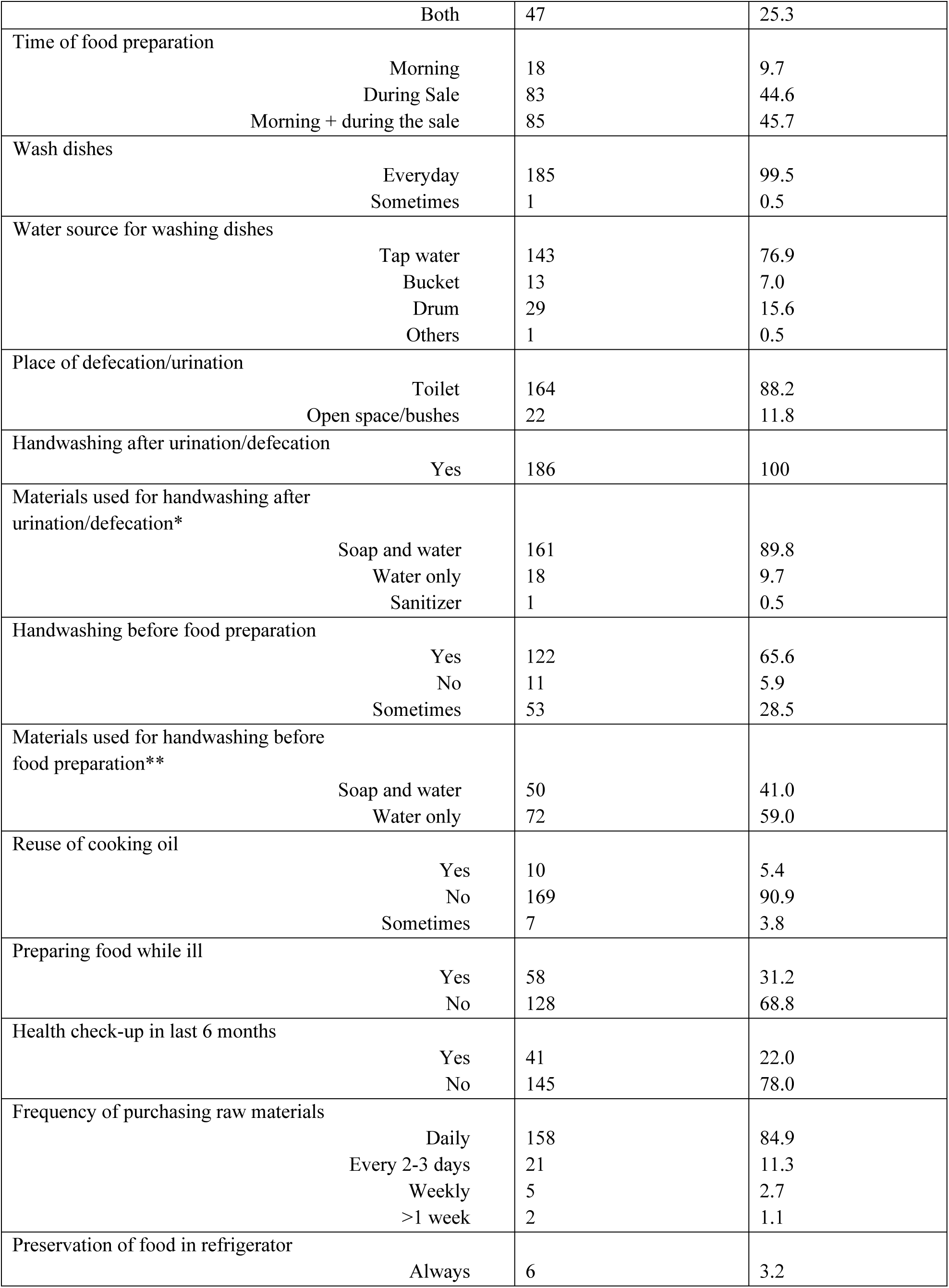
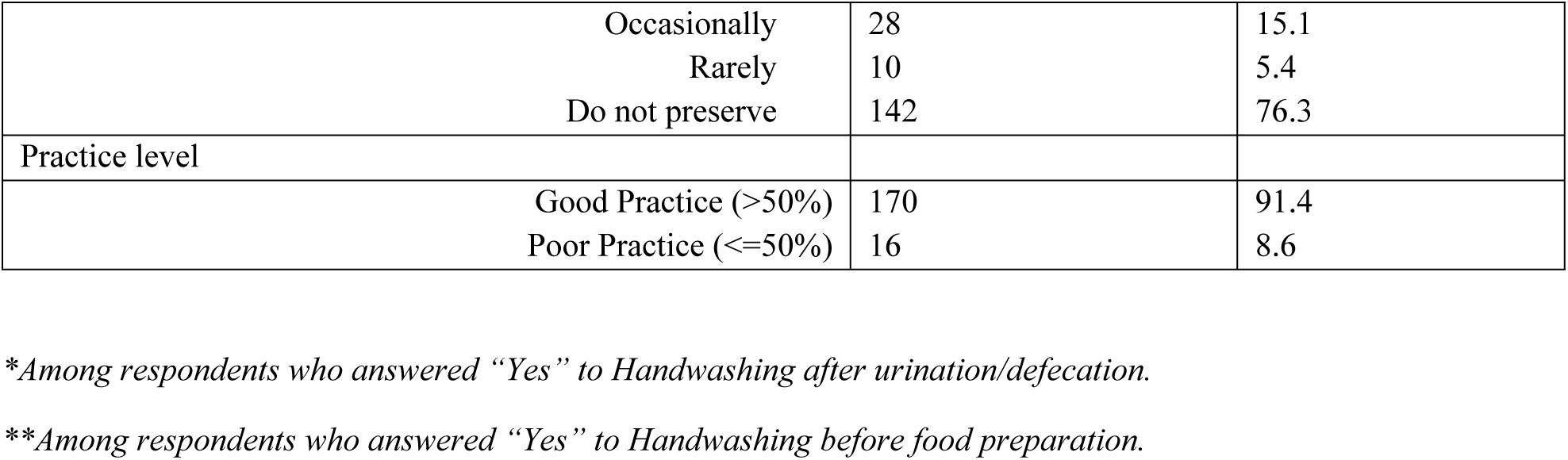
Type and preparation of food in the vending unit (N=186)

| Characteristics | Frequency | Percentage (%) |
| --- | --- | --- |
| Type of food (Multiple response) |  |  |
| Freshly cooked food | 154 | 82.8 |
| Pre-prepared food | 61 | 32.8 |
| Raw food | 8 | 4.3 |
| Place of food preparation |  |  |
| At home | 16 | 8.6 |
| At the vending site/marketplace | 123 | 66.1 |

|  |  |  |  |
| --- | --- | --- | --- |
|  | Both | 47 | 25.3 |
| Time of food preparation |  |  |  |
|  | Morning | 18 | 9.7 |
|  | During Sale | 83 | 44.6 |
|  | Morning + during the sale | 85 | 45.7 |
| Wash dishes |  |  |  |
|  | Everyday | 185 | 99.5 |
|  | Sometimes | 1 | 0.5 |
| Water source for washing dishes |  |  |  |
|  | Tap water | 143 | 76.9 |
|  | Bucket | 13 | 7.0 |
|  | Drum | 29 | 15.6 |
|  | Others | 1 | 0.5 |
| Place of defecation/urination |  |  |  |
|  | Toilet | 164 | 88.2 |
|  | Open space/bushes | 22 | 11.8 |
| Handwashing after urination/defecation |  |  |  |
|  | Yes | 186 | 100 |
| Materials used for handwashing after urination/defecation* |  |  |  |
|  | Soap and water | 161 | 89.8 |
|  | Water only | 18 | 9.7 |
|  | Sanitizer | 1 | 0.5 |
| Handwashing before food preparation |  |  |  |
|  | Yes | 122 | 65.6 |
|  | No | 11 | 5.9 |
|  | Sometimes | 53 | 28.5 |
| Materials used for handwashing before food preparation** |  |  |  |
|  | Soap and water | 50 | 41.0 |
|  | Water only | 72 | 59.0 |
| Reuse of cooking oil |  |  |  |
|  | Yes | 10 | 5.4 |
|  | No | 169 | 90.9 |
|  | Sometimes | 7 | 3.8 |
| Preparing food while ill |  |  |  |
|  | Yes | 58 | 31.2 |
|  | No | 128 | 68.8 |
| Health check-up in last 6 months |  |  |  |
|  | Yes | 41 | 22.0 |
|  | No | 145 | 78.0 |
| Frequency of purchasing raw materials |  |  |  |
|  | Daily | 158 | 84.9 |
|  | Every 2-3 days | 21 | 11.3 |
|  | Weekly | 5 | 2.7 |
|  | >1 week | 2 | 1.1 |
| Preservation of food in refrigerator |  |  |  |
|  | Always | 6 | 3.2 |
| Occasionally | 28 | 15.1 |
| Rarely | 10 | 5.4 |
| Do not preserve | 142 | 76.3 |
| Practice level |  |  |
| Good Practice (>50%) | 170 | 91.4 |
| Poor Practice (<=50%) | 16 | 8.6 |

*\*Among respondents who answered “Yes” to Handwashing after urination/defecation.*
*\*\*Among respondents who answered “Yes” to Handwashing before food preparation.*

**Table 4:**
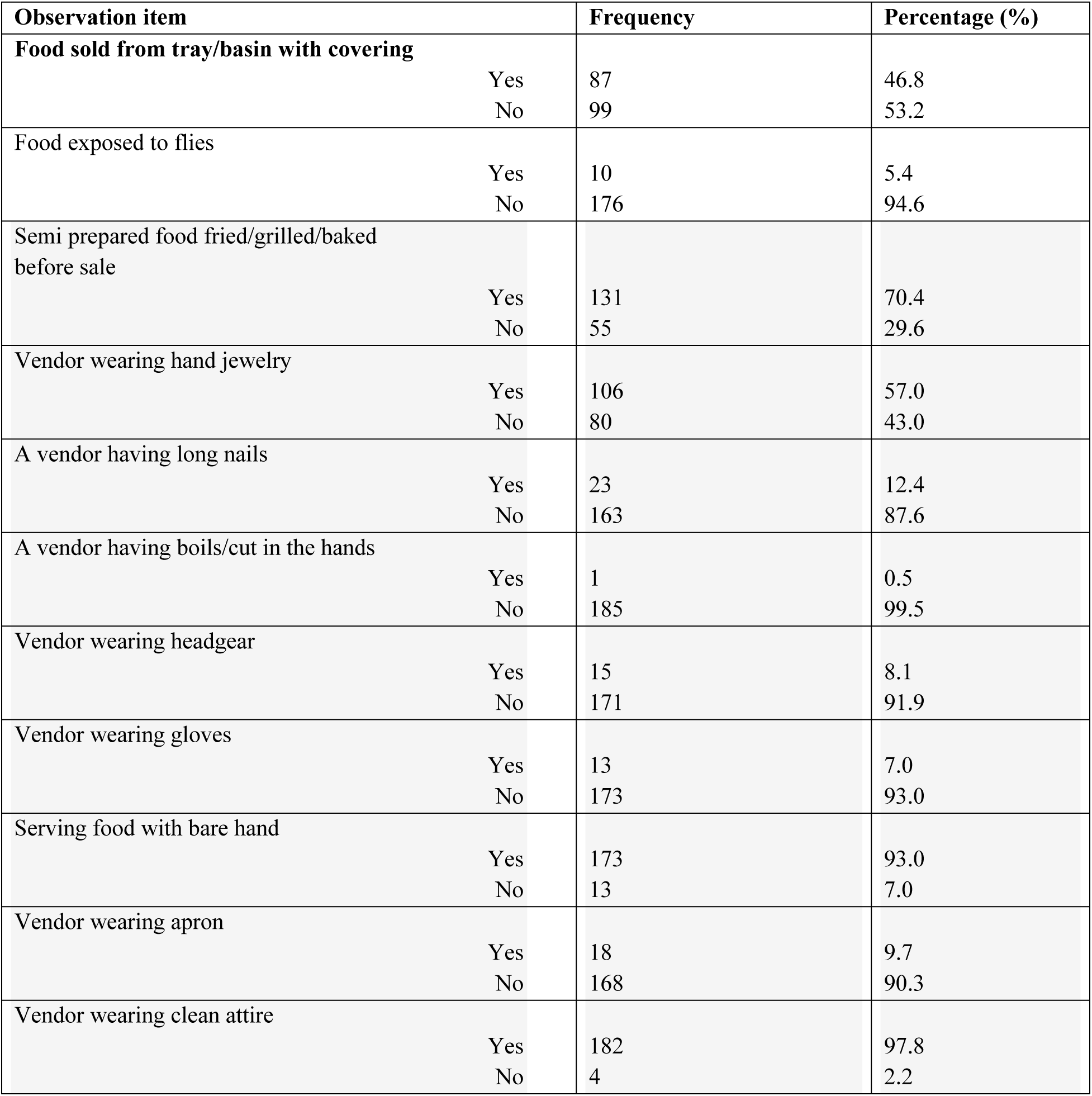
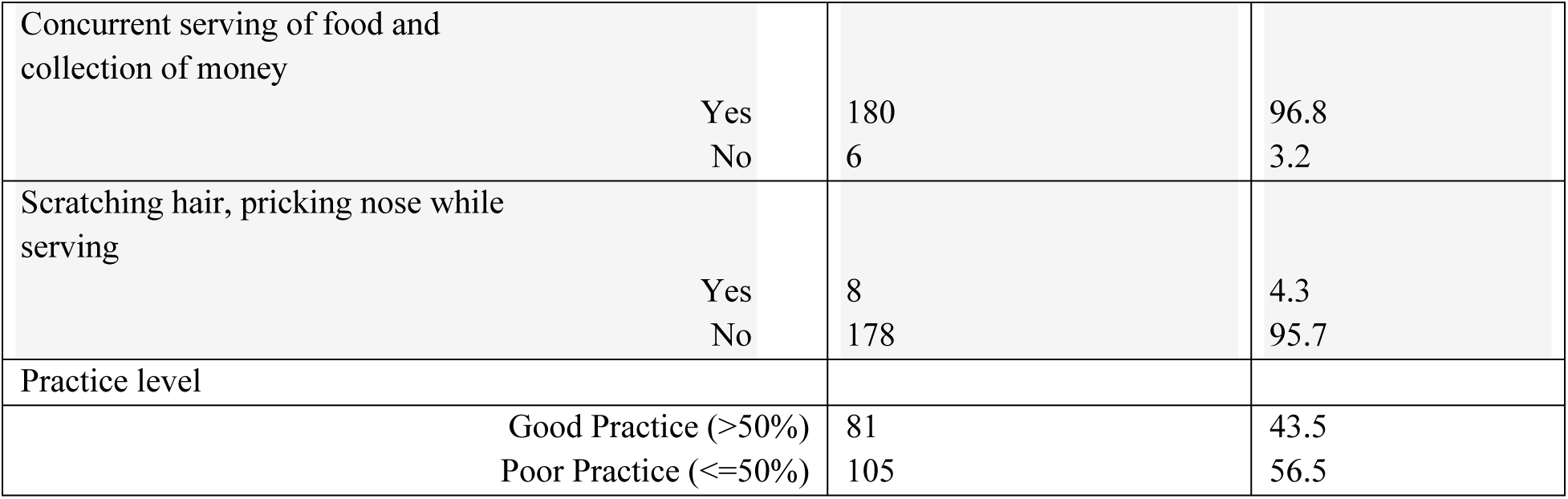
Observed food handling practices(N=186)

| Observation item | Frequency | Percentage (%) |
| --- | --- | --- |
| **Food sold from tray/basin with covering** |  |  |
| Yes | 87 | 46.8 |
| No | 99 | 53.2 |
| **Food exposed to flies** |  |  |
| Yes | 10 | 5.4 |
| No | 176 | 94.6 |
| **Semi prepared food fried/grilled/baked before sale** |  |  |
| Yes | 131 | 70.4 |
| No | 55 | 29.6 |
| **Vendor wearing hand jewelry** |  |  |
| Yes | 106 | 57.0 |
| No | 80 | 43.0 |
| **A vendor having long nails** |  |  |
| Yes | 23 | 12.4 |
| No | 163 | 87.6 |
| **A vendor having boils/cut in the hands** |  |  |
| Yes | 1 | 0.5 |
| No | 185 | 99.5 |
| **Vendor wearing headgear** |  |  |
| Yes | 15 | 8.1 |
| No | 171 | 91.9 |
| **Vendor wearing gloves** |  |  |
| Yes | 13 | 7.0 |
| No | 173 | 93.0 |
| **Serving food with bare hand** |  |  |
| Yes | 173 | 93.0 |
| No | 13 | 7.0 |
| **Vendor wearing apron** |  |  |
| Yes | 18 | 9.7 |
| No | 168 | 90.3 |
| **Vendor wearing clean attire** |  |  |
| Yes | 182 | 97.8 |
| No | 4 | 2.2 |

|  |  |  |  |
| --- | --- | --- | --- |
| Concurrent serving of food and collection of money | Yes<br>No | 180<br>6 | 96.8<br>3.2 |
| Scratching hair, pricking nose while serving | Yes<br>No | 8<br>178 | 4.3<br>95.7 |
| Practice level |  |  |  |
| Good Practice (>50%) |  | 81 | 43.5 |
| Poor Practice (<=50%) |  | 105 | 56.5 |

**Table 5:** Observed sanitary condition of vending site(N=186)

| Observation item |  | Frequency | Percentage (%) |
| --- | --- | --- | --- |
| Vending site located on or near a clean and well-maintained road | Yes<br>No | 174<br>12 | 93.5<br>6.5 |
| Presence of waste bin | Yes<br>No | 174<br>12 | 93.5<br>6.5 |
| Presence of refuse dump site nearby vending site | Yes<br>No | 30<br>156 | 16.1<br>83.9 |
| Presence of hand towel | Yes<br>No | 134<br>52 | 72.0<br>28.0 |
| Presence of soap for hand-washing | Yes<br>No | 92<br>94 | 49.5<br>50.5 |
| Presence of rats/cockroaches | Yes<br>No | 2<br>184 | 1.1<br>98.9 |
| Washing utensil with soap and running water | Yes<br>No | 167<br>19 | 89.8<br>10.2 |
| Clean water collector or water disposal | Yes<br>No | 150<br>36 | 80.6<br>19.4 |
| Presence of non-porous surface | Yes<br>No | 101<br>85 | 54.3<br>45.7 |
| Presence of Latrine Nearby |  |  |  |
|  | Yes | 161 | 86.6 |
|  | No | 25 | 13.4 |
| Sanitary condition |  |  |  |
|  | Good (>50%) | 176 | 94.6 |
|  | Poor (<=50%) | 10 | 5.4 |

**Table 6:** Association between sociodemographic factors and food hygiene knowledge level.

| Variables | Knowledge category |  | χ<sup>2</sup> or Fisher's | df | P value | OR (95% CI) |
| --- | --- | --- | --- | --- | --- | --- |
|  | Poor Knowledge | Good Knowledge |  |  |  |  |
| Sex of Respondents |  |  |  |  |  |  |
| Male | 55 (56.1%) | 43 (43.9%) | 1.862 | 1 | 0.172 | - |
| Female | 58 (65.9%) | 30 (34.1%) |  |  |  |  |
| Education level of Respondents |  |  |  |  |  |  |
| Illiterate | 49 (79%) | 13 (21%) | 13.033 | 1 | <0.001 | 3.534 (CI 1.745 – 7.156) |
| Literate | 64 (51.6%) | 60 (48.4%) |  |  |  |  |
|  | 104 (60.4%) | 68 (39.6%) | 0.079 | 1 | 0.778 | - |
|  | 9 (64.2%) | 5 (35.7%) |  |  |  |  |
| Type of food vending |  |  |  |  |  |  |
| Fixed stall |  | 25 (36.8%) | 0.277 | 1 | 0.599 | - |
| Mobile |  | 48 (40.7%) |  |  |  |  |
| Years of working |  |  |  |  |  |  |
| ≤ 5 years | 54 (58.8%) | 36 (41.2%) | 0.041 | 1 | 0.839 | - |
| >5 years | 59 (61.4%) | 37 (38.6%) |  |  |  |  |
| Training on food safety |  |  |  |  |  |  |
| Yes | 0 (0%) | 1 (100%) | N/A | - | - | - |
| No | 113 (61.1%) | 72 (38.1%) |  |  |  |  |

*The Mann-Whitney U test showed no statistical significance in age and food hygiene knowledge level (p = 0.660).*

**Table 7:**
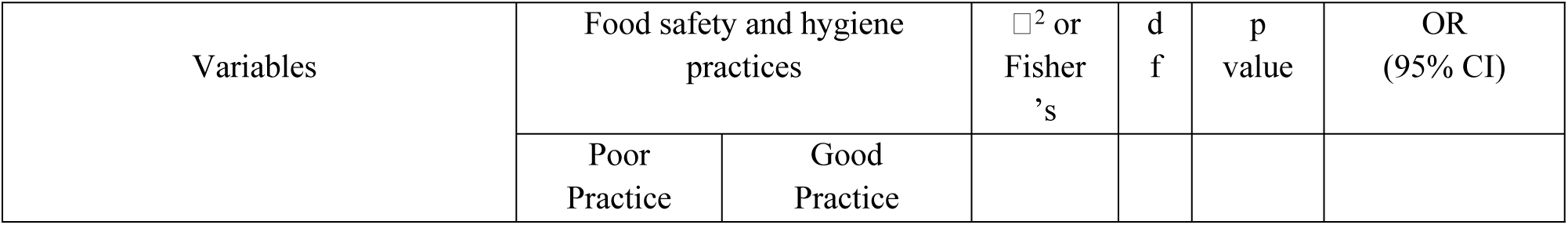
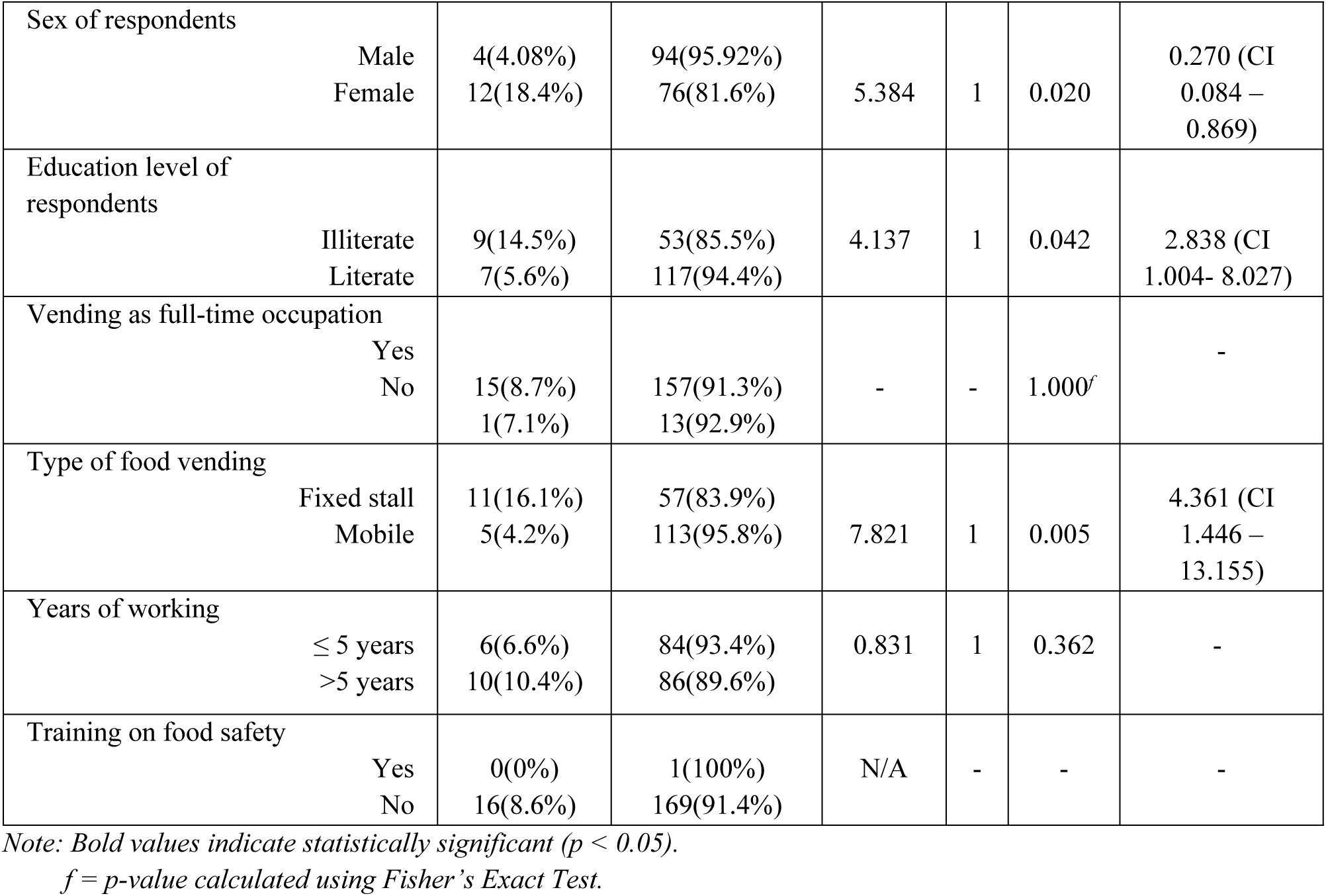
Association between sociodemographic factors and food safety and hygiene practice level.

| Variables | Food safety and hygiene practices |  | χ<sup>2</sup> or Fisher's | df | p value | OR (95% CI) |
| --- | --- | --- | --- | --- | --- | --- |
|  | Poor Practice | Good Practice |  |  |  |  |

|  |  |  |  |  |  |  |  |
| --- | --- | --- | --- | --- | --- | --- | --- |
| Sex of respondents | Male<br>Female | 4(4.08%)<br>12(18.4%) | 94(95.92%)<br>76(81.6%) | 5.384 | 1 | 0.020 | 0.270 (CI<br>0.084 –<br>0.869) |
| Education level of respondents | Illiterate<br>Literate | 9(14.5%)<br>7(5.6%) | 53(85.5%)<br>117(94.4%) | 4.137 | 1 | 0.042 | 2.838 (CI<br>1.004- 8.027) |
| Vending as full-time occupation | Yes<br>No | 15(8.7%)<br>1(7.1%) | 157(91.3%)<br>13(92.9%) | - | - | 1.000<sup>f</sup> | - |
| Type of food vending | Fixed stall<br>Mobile | 11(16.1%)<br>5(4.2%) | 57(83.9%)<br>113(95.8%) | 7.821 | 1 | 0.005 | 4.361 (CI<br>1.446 –<br>13.155) |
| Years of working | ≤ 5 years<br>>5 years | 6(6.6%)<br>10(10.4%) | 84(93.4%)<br>86(89.6%) | 0.831 | 1 | 0.362 | - |
| Training on food safety | Yes<br>No | 0(0%)<br>16(8.6%) | 1(100%)<br>169(91.4%) | N/A | - | - | - |

*Note: Bold values indicate statistically significant ( $p < 0.05$ ).*
*f = p-value calculated using Fisher's Exact Test.*

**Table 8:** Association between food hygiene knowledge level and food safety and hygiene practice level.

| Knowledge level | Food safety and hygiene practice |  | X<sup>2</sup> | df | P value | OR (95% CI) |
| --- | --- | --- | --- | --- | --- | --- |
|  | Poor Practice | Good Practice |  |  |  |  |
| Poor Knowledge | 12(10.7%) | 100(99.3%) | 1.490 | 1 | 0.222 | - |
| Good Knowledge | 4(5.5%) | 69(94.5%) |  |  |  |  |

## Discussion

This study describes the demographic status of street food vendors, their knowledge level, food safety and hygiene practice level, observed food handling practices, and sanitation conditions of vending sites, and the association between sociodemographic characteristics, knowledge, and practices regarding food hygiene and safety.

The median age of the respondents was 38.50 years (IQR: 29.75–46.25). This indicates that the productive age group works as street food vendors, which is like the studies from low- and middle-income countries. (16) Male vendors were slightly more than female vendors which shows alignment with a similar study done in Kathmandu city. (17)

Majority of them had educational status as primary level or less(70.9%), which is in line with the findings from Dhading, Nepal, and Bangladesh which report that lack of knowledge is a key driver unhygienic food practices among food vendors. (2,7)

Most of the food vendors worked full-time as food venders, similar to findings from the study done in Kaski. (3)The majority of the food vendors were mobile vendors, which contradicts the findings from a study conducted in southwestern Nigeria. (8,18) Over half of the food sellers have been in this profession for more than five years, according to the report. Most of the Street food vendors received no formal food safety training. Similar results were seen in other studies, where most of the street food vendors lacked training in food safety. (4,19) This points towards possible regulatory actions required from government.

Studies from Bangladesh, Ethiopia and Nepal show similar findings. Research from Bangladesh, Ethiopia and Nepal supports this conclusion. For example, research from Bangladesh, Ethiopia and Nepal found that street food vendors need to learn more about food **hygiene.** Most of the 74 street food vendors said they recognized signs of food poisoning like diarrhea, vomiting, nausea and abdominal pain in this study. Knowledge regarding food borne illnesses is necessary in order to improve wash and food related safety pratices. The study demonstrated that being a male or a female street vendor has no significant effect on the level of knowledge regarding food hygiene, supporting the results of a study conducted in another regions of Nepal. (7)

However, the educational status of the respondents was significantly found to be associated with the knowledge level on food hygiene (◻^2^ = 3.033, df = 1, p < 0.001), which is consistent with the findings from the study done in Dhading, Nepal Chattogram City, Bangladesh and the Metropolitan District in Ghana. (6) The odds of having good knowledge were 3.5 times higher among literate street food vendors than among illiterate ones (OR = 3.53). Food hygiene knowledge was not significantly associated with full-time food vending, indicating that full-time food vending may not be sufficient to improve food hygiene knowledge. This finding further emphasizes how crucial food hygiene education and training programmes are to improving food hygiene awareness. It was found that any type of street food vending doesn’t make a significant association with the knowledge level of food hygiene in this study, which is consistent with the findings from a study conducted in Gondar City, Northwest. (12)

The study shows that there is no significant association between food hygiene knowledge and experience or working years. The Mann-Whitney U test was conducted to compare the median age of street food vendors with food hygiene knowledge, as age was not normally distributed. However, findings indicated that no significant difference in the median age of vendors with good knowledge and poor knowledge. (U = 3967.00, Z = -0.440, p = 0.660). This shows that age is not a determining factor for food hygiene knowledge among street food vendors, similar to the findings from the studies done in Bangladesh and Ethiopia. (11)

Though vendors had poor knowledge on food hygiene, the majority of them had good self-reported food safety and hygiene practices which cpuld be as result of desirability bias in the study. The result revealed that there is a significant association between food safety and hygiene practices with gender (◻^2^ = 5.384, df = 1, p = 0.020).

Additionally, there was a significant association between education level and food safety and hygiene practices (◻^2^ = 4.137, df = 1, p = 0.042). This suggests that education may help people in adopting hygienic behaviours. But, food vending as a full-time occupation didn’t show association with food safety and hygiene practices supporting a study done in Ghanaindicating that continuous involvement in food vending doesn’t ensure good hygienic behaviour of the street food vendors.(18,20)

A significant association was observed between type of food vending and food safety practices (◻^2^ = 7.821, p = 0.005). Vendors operating from fixed stalls are more than four times likely as to exhibit poor food safety and hygiene practices (OR = 4.361, 95% CI: 1.446 – 13.155). The study showed no association between working experience and food safety and hygiene practices, opposing the findings from the study done in Ghana. (19) Mann-Whitney U test was used to compare the median age of street food vendors with good and poor food safety and hygiene practices. The result showed that there is no statistically significant difference in median age between vendors with food hygiene and safety practices level (U = 1059.50, Z = -1.460, p = 0.144), suggesting age doesn’t significantly influence food safety and hygiene practices among street food vendors.

Association was not observed between overall knowledge level and overall food safety practice. Which shows that having knowledge doesn’t necessarily lead to good practices; factors like infrastructure and regulation may play a crucial role in food hygiene practice. This finding is consistent with study from South Africa and low-middle-countries, which reported that knowledge alone doesn’t ensure good food hygiene practice. (21)This further emphasizes that reinforcing and motivational factors must be put into action in addition to knowledge and awareness to bring change in actual vending pratices.(22)

From the study, there is no association between knowledge and type of food prepared and also a no association were observed between food hygiene knowledge and food preparation place, whether at home or at a vending site or both. However, the significant association was seen between food hygiene knowledge and food preparation time (◻^2^ = 8.590, df = 2, p = 0.014).

Most of the street food vendors washed their dishes daily whether they had good knowledge or poor knowledge. Source of water for washing dishes and place of defecation / urination were not significantly associated with knowledge level. Which suggests that these practices are influenced by availability of infrastructure rather than awareness about hygiene. Handwashing after defecation/ urination was done by all the respondents. Materials used for handwashing after defecation / urination and before food preparation were not found associated with food hygiene knowledge which supports the findings from Ethiopia and Ghana which explains that access to soap and sanitizer is related to financial condition rather than knowledge. (2) Similarly, reuse of cooking oil was not significantly associated with the knowledge of the vendors. Preparation of food while ill was not significantly associated with the knowledge of the vendor. Studies have shown that street food vendors may continue preparing food while being ill because of economic pressure. Health checkup status in last six months was not significantly associated with food hygiene knowledge. This shows that those having food hygiene knowledge may positively affect the health-seeking behavior. Majority of the street food vendors reported that they purchased materials everyday as per daily requirement rather than selling stale items. (23)

However, observation based findings revealed different results in varying degrees than self-reported data. We observed that more than half of the vendors sold food without covering. They reported that people get attracted to their stalls if they see ready-made foods. Despite of food being sold from the tray without covering, the food was least exposed to the flies. This might be due to seasonal variation leading to least active flies in environment as the data collection was carrieed out during winter season.(24)

The use of protective equipment like gloves, headgear, and apron was low, while vendors wearing hand jewelry was higher posing threat to food contamination. Most of the street food vendors were wearing clean attire and had short nails. We observed that most of the street food vendors kept semi-prepared food in the stall, reporting that it saves time for preparing food. Concurrently serving and collecting money was observed in most of the vendors. With no additional staffs at the stall, vendors have to perform multiple tasks while serving the customers. On observation, almost all vendors hands were injury/boils free. More than half of the vendors demonstrated poor food handling practices based on observation scores despite reporting good hygiene practices. This may be due to social desirability bias in self-reported data and provides emphasis on direct observation in food safety assessment.(4,25)

The sanitary condition of the vending sites was also observed and generally found satisfactory. Most of the stalls were located near the clean roads, had waste bins, access to latrines, clean water sources, and almost an absence of pests. These findings show good environmental conditions for street food vendors to practice good food vending. However, an absence of soap at nearly half of the vendors and having nonporous surfaces make pose a risk for foodborne disease transmission.

Overall, the findings indicate a gap in knowledge and practice. Knowledge alone can’t lead to improved hygiene practices. Multiple factors like training, infrastructure, supervision, and strict enforcement of food safety regulation are necessary.

### Recommendations

- Regular food safety and hygiene training program should be given for street food vendors.

- Licensing and regular inspection should be done to ensure food hygiene standards.

- Shifting vendors to places were infrastructures like handwashing facility, toilet etc. is available.

- Periodic health check-ups for street food vendors should be encouraged.

- A food safety promotion program should be organized.

- Vendors should be encouraged to follow safe food handling practices including use of protective equipment and avoiding food preparation while ill.

### Limitations of the study

The cross-sectional design limits the causal inferences. Seasonal variation may have affected observation findings. Data collection through face-to-face interviews may have been subject to social desirability bias, where participants potentially overstated good hygiene practices. Therefore we have tried to minimize the bias using an observational practice to see actual practice. However, we cannot confirm that these observational checklist can be relied upon for their everyday practices. The study focused only on the vendors’ knowledge and practices, excluding consumer experiences. Microbiological testing was not conducted.

## Conclusion

This study assessed food hygiene knowledge, food safety and hygiene practices, and associated factors among street food vendors of Chitwan and Makawanpur Districts of Bagmati Province, Nepal. The findings indicate that street food vending is the occupation mainly done by individuals of productive age group. Less than half of the vendors showed good food hygiene knowledge indicating gap in knowledge. Education was found to be significantly associated with food hygiene knowledge however, gender, age, working experience, type of vending and full-time engagement in food vending were not significantly associated with knowledge level. Despite having low level of knowledge, majority of the vendors reported good safety and hygiene practices. Gender, education and type of food vending was significantly associated with food safety and hygiene practices while age, work experience and full-time vending were not significantly associated with practice. There was no significant association between overall knowledge and overall practices indicating knowledge only does not lead to safe food hygiene practices. Observation revealed poor food handling practices. The sanitation condition of most vending sites was satisfactory.

The study contributes to evidence by showing that structural and contextual factors, such as access to water supplies, sanitation facilities, and monitoring, are important determinants of food hygiene practices among street food vendors. These results highlight the necessity of comprehensive food safety initiatives that go beyond raising awareness. The study has many limitations in spite of its contributions. Causal inference between related factors and hygiene practices is limited by the cross-sectional methodology. Seasonal fluctuation may have impacted observational data, and social desirability bias may have impacted self-reported habits. Furthermore, it is more difficult to objectively determine the degree of food contamination when microbiological testing is not conducted. Overall, the study shows a significant discrepancy between street food vendors practices, and knowledge of food hygiene. To enhance food hygiene, awareness and capacity building along with provision of supportive infrastructure, effective legislation, regular inspections of vending sites should be done.

## Declarations

## Acknowledgement

We would like to acknowledge the faculties of department of Public Health of Madan Bhandari Academy of Health Sciences who provided their support in completion of this research project. Also, we would extend our sincere thanks to University grants commission who allocated student support funding to the supervisor of this study.

## Data Availability Statement

Data will be made available upon request to the authors via email.

